# The Health Condition Timeline as a Model for Pregnancy Disease Management

**DOI:** 10.1101/2023.02.06.23285418

**Authors:** Scott McLachlan, Bridget J Daley, Kudakwashe Dube, Evangelia Kyrimi, Martin Neil, Norman E Fenton

## Abstract

Process flow diagrams like caremaps are common in clinical practice guidelines and treatment texts. However, their context is often limited to a single diagnostic or treatment event. While a method has been proposed for creating a health and disease lifecycle called the *health condition timeline* (HCT), that method is yet to be demonstrated for an entire health condition. This paper investigates development of an HCT for *gestational diabetes mellitus* (GDM), and whether the HCT and caremaps it incorporates can be used to support patient care to develop decision support tools. We show that this approach can be used to expedite development of clinical decision-support and clinician- and patient-facing applications. Caremaps, HCT and the decision support tools created with them could improve patient awareness for their condition and reduce the impact of their disease on themselves and the limited resources of our healthcare systems.

## 1. Introduction

Caremaps are graphical representations of the sequence of patient care activities for a medical condition [1]. Caremaps have existed in various forms since the 1980’s, and contemporary caremaps are often seen in clinical practice guidelines [1]. Caremaps have been used to describe discrete portions of the overall care of individuals with such chronic conditions as diabetes. Examples include compact models describing care for diabetic dietary issues [2], a simplified 6-node caremap of the gross management phases for treatment of hyperglycaemia in patients taking atypical antipsychotic medications [3], and a comprehensive model of the pathways and decision points for the labour and birth event [4]. Caremaps have been used to visually describe the expected surgical care process to maternity patients who are going to have, or have had, a caesarean [5], and they have been populated with statistics and probabilities for use in creating *realistic synthetic electronic health records* (RS-EHR) and health AI tools [6, 7]. However, caremaps and RS-EHR still only consider singular disease or treatment incidents. Recently, the caremap and RS-EHR models were extended to incorporate an overarching timeline structure for representing whole-of-life health and disease known as the *health condition timeline* (HCT) [8]. However, there are currently no examples in the literature of caremaps that describe the entire pregnancy event, or the diagnosis and management of an entire pregnancy impacted by pregnancy-related disease such as *gestational diabetes mellitus* (GDM). This is the term used where diabetes’ onset or first diagnosis occurs during pregnancy.

Process and timeline-based information visualisations (IV) have been used in management and evaluation of health service procedures, efficiency, safety and quality of care [19–23]. Among other purposes, caremaps have been used to coordinate care and aid clinicians and patients to anticipate activities along the pathway of disease, treatment and patient care [24–26]. A key difference is that prior caremap-based solutions generally focused on a single activity or component from a much larger care process, while the HCT solution describes the entire care process as a complete set of interconnected caremaps. Sequential task or activity-oriented IV approaches are often used to improve situational awareness and decision-making [19, 27, 28]; to understand clinical guideline and care process adherence and variation [19]; to visualise a timeline of care activities from raw patient data [29]; and to model the flow of activities in community nursing care [27]. In clinical education and patient care these IV can aid identification and articulation of relationships between behavioural and psychological symptoms [30] and improve clinical perception by visually organising patient care or clinical decision-making pathways [31, 32]. They have also been used to generate privacy preserving synthetic patient records for use in clinical education, health systems development [7], and development of clinical artificial intelligence (AI) [33].

Diabetes is the most common metabolic disorder of pregnancy and carries multiple risks for both mother and baby [9, 10]. Depending on an array of factors and the diagnostic criteria used, GDM occurs in between 2% - 25% of pregnancies. Incidence in the United Kingdom (UK) can be as high as 17% [12–14]. Between 40% - 60% of women diagnosed with GDM will go on to develop *Type 2 Diabetes Mellitus* (T2DM) later in life, and women with GDM are more likely to require caesarean delivery [11]. In a previous study members of this team evaluated the methodological quality of *clinical practice guidelines* (CPG) for GDM [11]. This work builds on that study by investigating development of a *health condition timeline* (HCT) for GDM that is consistent with those CPG; incorporating caremaps representing the diagnosis, initial treatment, delivery decisions and other essential phases that arise when managing GDM for the pregnant woman.

## 2. Background

The HCT can be constructed to represent the lifecycle of health and disease from two perspectives:

1. the individual patient’s perspective - describing the flow of health and disease experienced from birth to (eventual) death [8];
2. the perspective of the disease of interest - describing the flow from symptom identification and diagnosis through the care model and entire gamut of treatment options, and concluding with known health outcomes.

Our original work introducing HCT presented a solution focused solely on the first or *whole of life* perspective that generates realistic synthetic electronic health records (RS-EHR) for use in secondary research, health system development and clinical training [8]. We now re-visit HCT and propose a novel solution for using HCT from the second or *disease lifecycle* perspective.

This work was undertaken as part of an EPSRC-funded interdisciplinary collaboration between clinical researchers and computing and decision scientists to aid people with conditions like GDM. The PamBayesian project’s goal was to develop tools that offer real-time monitoring and clinical decision support in the community setting in order to increase patient independence and decrease reliance on direct consultation (www.pambayesian.org).

## 3. Method

The PamBayesian approach involved: (i) reviewing CPGs and literature relevant to the disease of interest; (ii) expert elicitation to provide broad and contextual understanding of the medical condition, treatment and potential outcomes; (iii) developing caremaps that describe the process, clinical decision points and potential outcomes; (iv) identifying idioms that describe common components of the caremap; (v) developing causal structures such as Bayesian Networks (BN) and Inference Diagrams (ID) based on the presence, order and interconnected nature of identified idioms; and (vi) paramaterising *conditional probability tables* (CPT) within the causal structure based on statistics from academic literature, or training from prospective and retrospective datasets of anonymised and/or aggregated patient records. Some case studies also incorporated an additional step that could be described as (iii-a) wherein every node and pathway of the caremap is populated with aggregate statistics regarding the patient population during a given period for a health provider or district so that clinical decision points and potential outcomes can be weighted and the caremap can be directly used in, or for development of, simple decision support. This work also required adding step (iii-b) to create the overarching HCT describing disease and treatment progression, in this example: from diagnosis of GDM through to postnatal follow-up with the woman six weeks after delivery of her baby. The complete methodology is shown in Figure 1.

**Figure 1:**
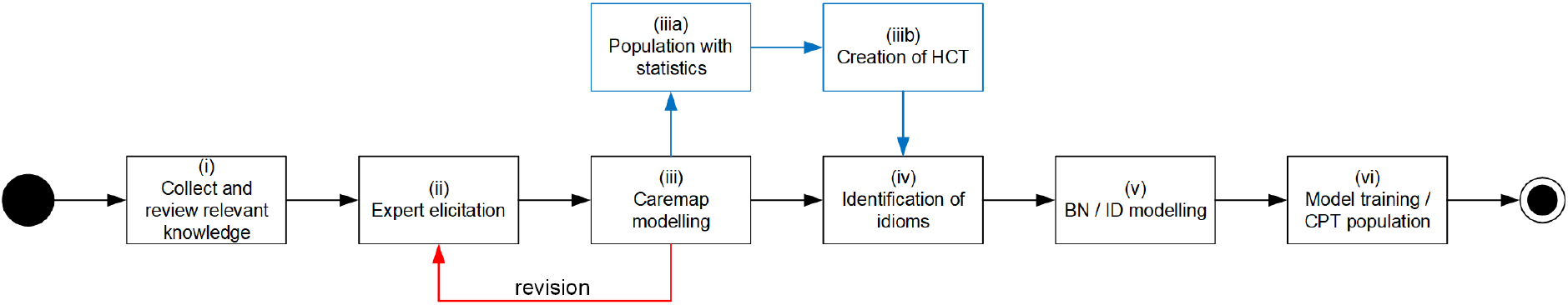
Methodology

## 4. Results

This section presents the overarching HCT for GDM and describes the individual component caremaps and their relationship to the NICE Guideline for Diabetes in Pregnancy (NICE, 2015). The HCT and caremaps were developed through collaboration between a midwife and a decision scientist whose primary focus was information visualisation. Caremaps were then presented to other obstetric and midwifery team members for at least two rounds of multidisciplinary review.

### 4.1 Health Condition Timeline for GDM

The HCT in Figure 2 is divided into four streams. The generic *patient stream* describes the linear pathway from symptom identification and diagnosis through treatment to the pregnancy outcome. It represents the period from conception of the baby through to postnatal discharge. The *clinical stream* describes the sequential pathway between different clinical events that arise either as part of natural disease progression or different treatment and management options. Each clinical event node represents an entire caremap and the arrows indicate the direction of flow between different nodes while managing the health condition. This component could be described as the *caremap of caremaps*. The *decision support stream* describes the overarching clinical decisions at each key stage of the health condition. Finally, the *communication and feedback* stream identifies key information that should be provided to the patient during that stage of the HCT.

**Figure 2:**
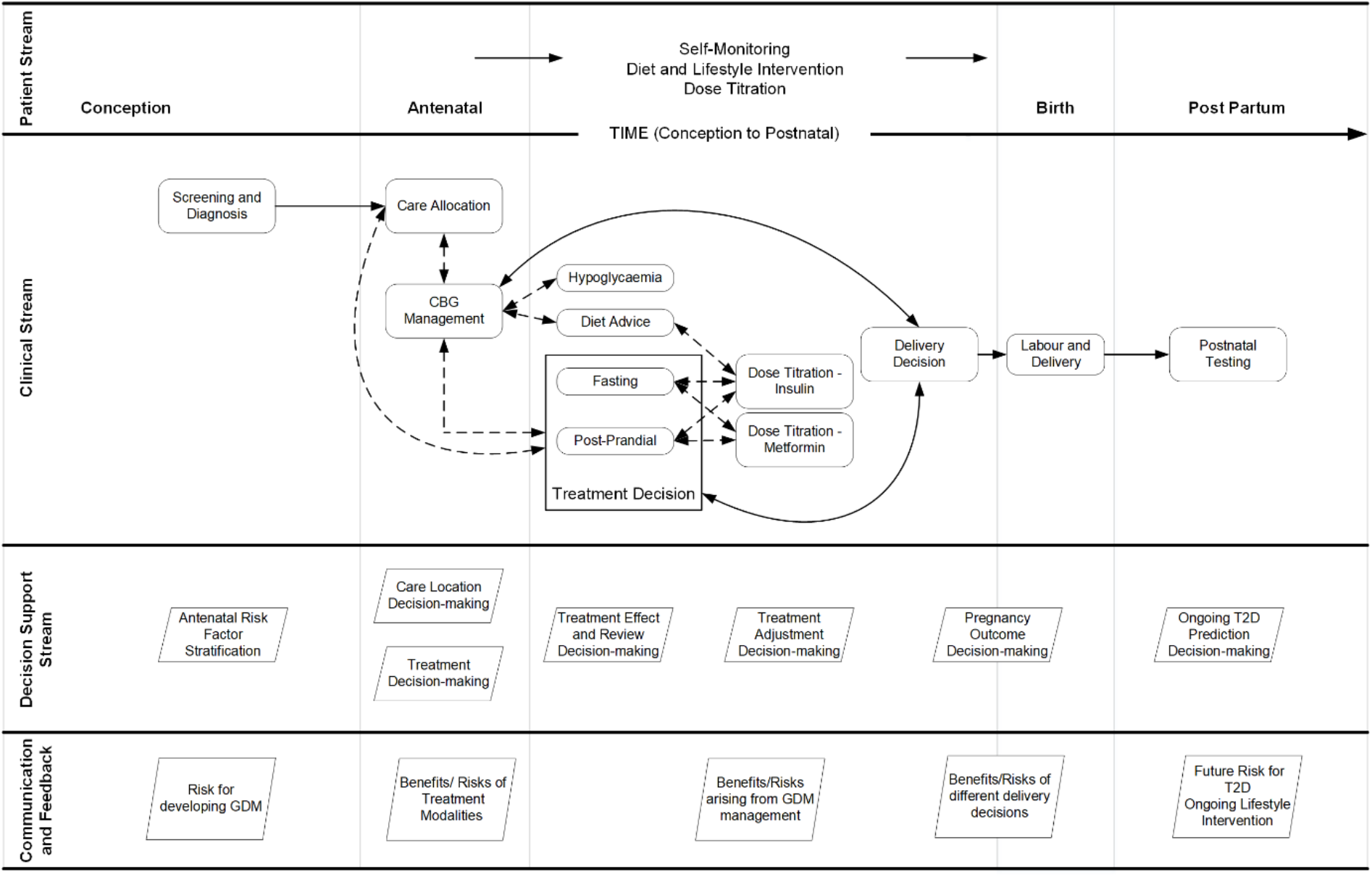
The Health Condition Timeline for GDM

### 4.2 Screening and Diagnosis

The Screening and Diagnosis phase is composed of two caremaps.

#### 4.2.1 Booking Visit

Caremap 1 describes the woman’s first attendance at the midwifery or obstetric clinic. It incorporates components from section 1.2 of the NICE guideline [15] that describe the risk assessment and necessary glucose testing processes. During the *booking visit*: (i) the pregnancy is confirmed using a *human chorionic gonadotropin* (HCG) urine test; (ii) the patient history and risk factors for GDM are collected; (iii) an initial clinical examination is performed, and (iv) if they have not already been performed by the GP, pregnancy-related blood tests are ordered. The standard clinical decisions arising during the booking visit are: (a) whether the woman has risk factors for GDM which necessitate an additional HbA1C test; and (b) verifying whether the pregnancy blood tests have already been performed in primary care and are available from the laboratory, EMIS, or a shared care record.

**Caremap 1.**
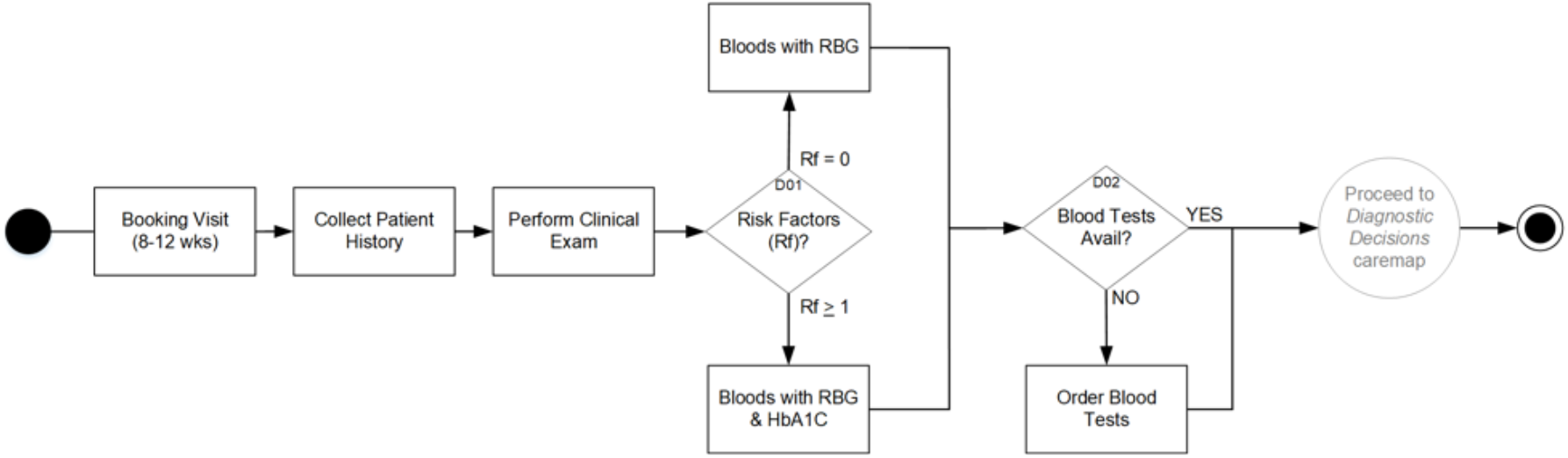
Booking Visit

#### 4.2.2 Diagnostic Decisions

Caremap 2 describes the primary assessment pathway. It diverges based on whether the pregnant woman was found to have risk factors for GDM which triggers an HbA1C test. It further diverges based on the results of that test. The incorporated testing and diagnosis processes arise out of sections 1.2.2 through 1.2.8 of the NICE guideline [15]. The diagnostic process classifies pregnant women into three distinct groups: (1) those who probably have pre-existing T2DM; (2) those diagnosed with GDM; and (3) those without GDM who will receive routine care.

**Caremap 2.**
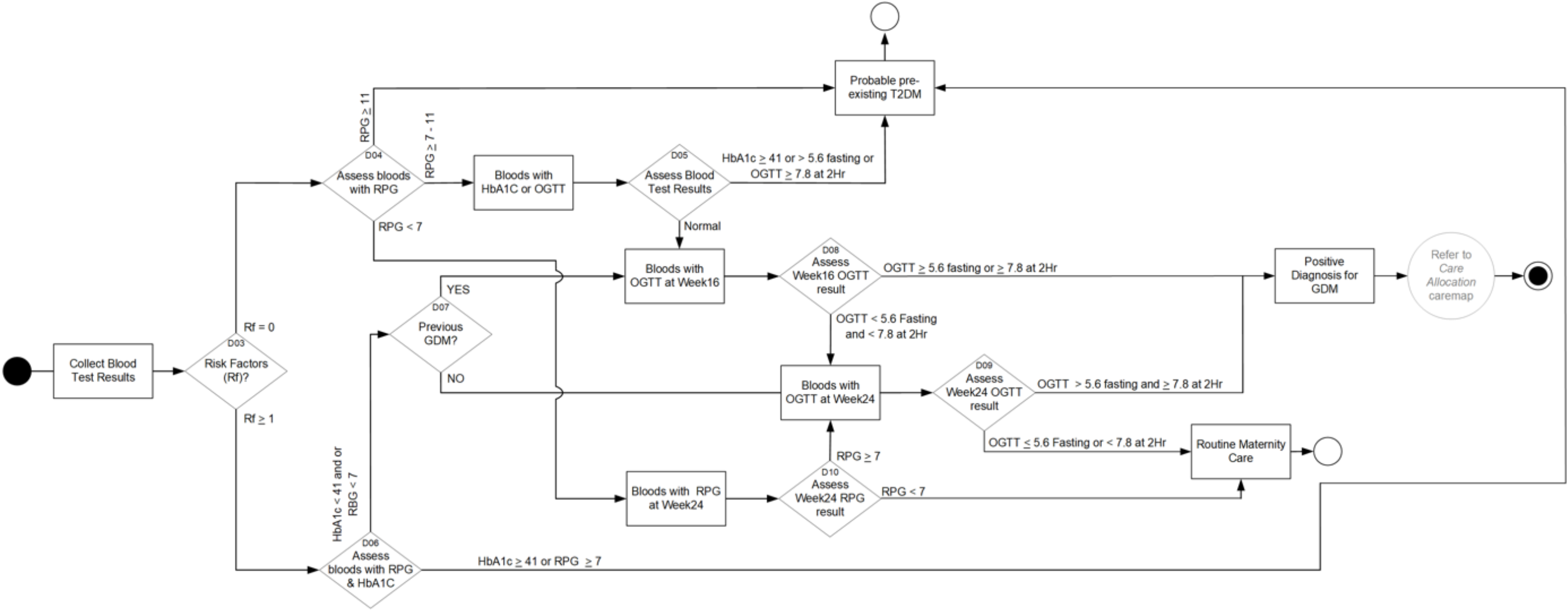
Diagnostic Decisions

### 4.3 Care Allocation

Caremap 3 is based on the review recommendations in section 1.2.9 and the antenatal care recommendations of section 1.3.37 - 1.3.38 of the NICE guideline [15] when deciding whether to assign the woman to primary care - *community midwifery care* (CMC), or secondary care - a hospital-based *diabetic antenatal clinic* (DANC). The decision is based on the degree which simple measures like diet control and gentle exercise (NICE sections 1.2.10 and 1.2.14 - 1.2.17) can maintain the woman’s GDM at sub-clinical levels. GDM control is assessed by self-administering and recording capillary blood glucose (CBG) tests several times each day.

**Caremap 3.**
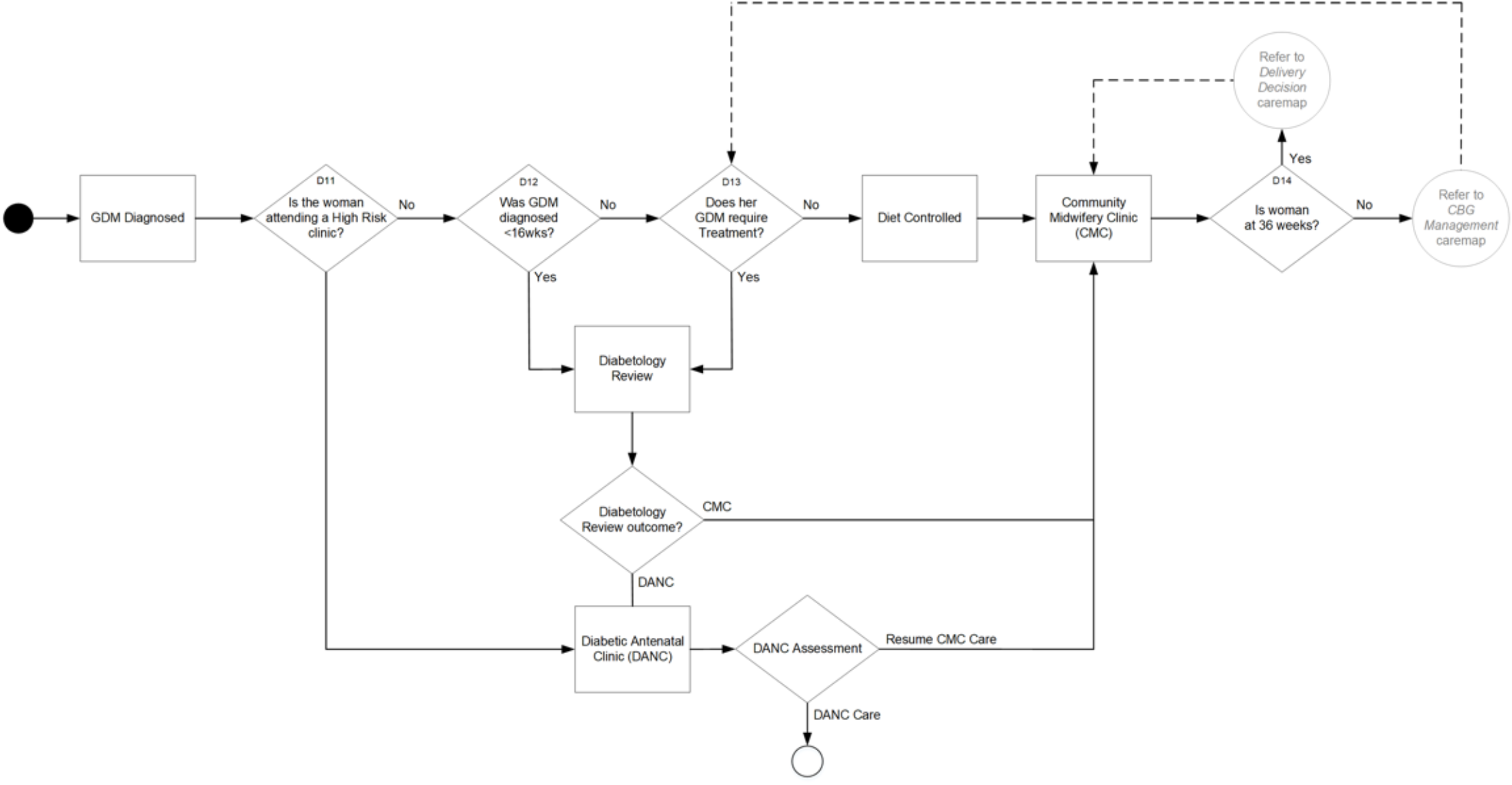
Care Allocation

### 4.4 CBG Management

After diagnosis of GDM we educate the woman in how to self-monitor her *capillary blood glucose* (CBG) (NICE section 1.2.11). Caremap 4 describes the pathway for managing GDM whether the woman presents as hypoglycaemic or hyperglycaemic. It informs and refers to other caremaps in the HCT, including the *delivery decision* caremap where the pregnant woman is at 36 weeks or later gestation, and the *treatment decisions* caremap where this is either the first instance of uncontrolled CBG, or where the clinician believes it will be difficult for the woman to manage her GDM without diabetic medication.

**Caremap 4.**
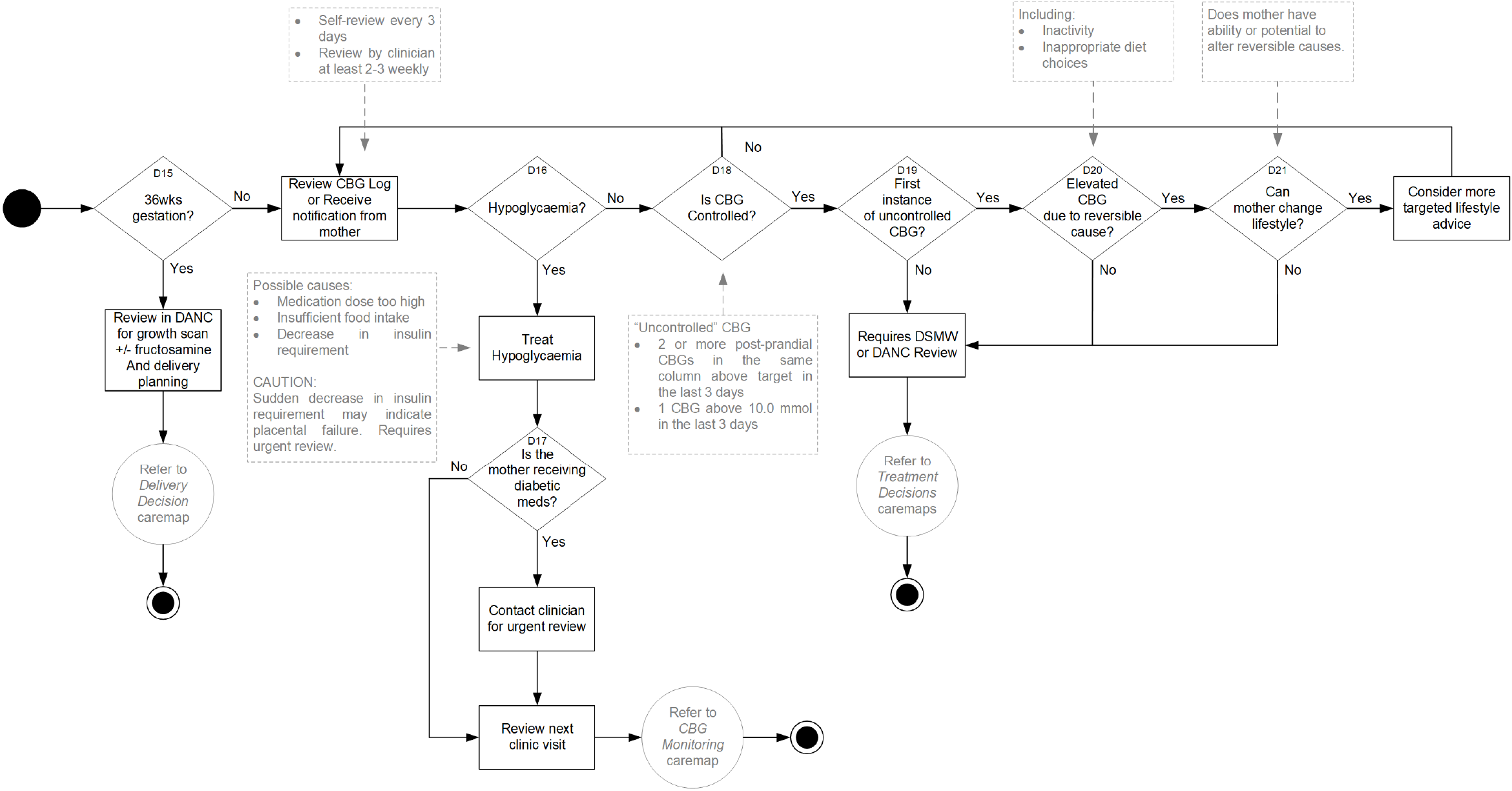
CBG Management

### 4.5 Treatment Decisions

There are two *treatment decisions* caremaps differentiated by the time of day the patient is testing her CBG.

#### 4.5.1 Treatment decision - Fasting

Caremap 5 incorporates the fasting test requirements of the NICE guideline at section 1.3 with the clinical levels described in sections 1.2.18 - 1.2.23 and 1.3.12 - 1.3.16. It considers GDM care based on the mother’s recent fasting (morning) CBG levels. During operation of this caremap that prescription of Metformin and Insulin will be considered. In each case, the chosen treatment will be reviewed in two weeks.

#### 4.5.2 Treatment Decision - Post-prandial

Caremap 6 incorporates the post-meal testing requirements of the NICE guideline at section 1.3 with the clinical levels described in sections 1.2.18 - 1.2.23 and

1.3.12 - 1.3.16. It considers GDM care based on the woman’s recent post prandial (after meal) CBG levels. During the operation of this caremap prescription of Metformin and Insulin will be considered. In each case, the chosen treatment will be reviewed in two weeks.

### 4.6 Hypoglycaemia

Caremap 7 describes a pathway the diabetic woman can follow after each CBG test to assess whether her glucose result or symptoms indicative of hypoglycaemia. This was developed to be patient-focused to demonstrate that caremaps: (a) can be approachable for the non-clinical lay person; and (b) could be used to develop algorithms, rules or software solutions for use by patients and their family or carers. This caremap reduces several pages of text contained in CPGs and patient handouts into a simple visual prompt. It presents a succession of decision points as questions (in the diamond-shaped nodes) and aids the pregnant woman in following the pathway to easily decide *what should I do next?* Even in situations where her hypoglycaemia is resolved, the caremap attempts to identify the underlying cause while still recommending that she contact her diabetic specialist midwife (DSMW) for advice.

**Caremap 5.**
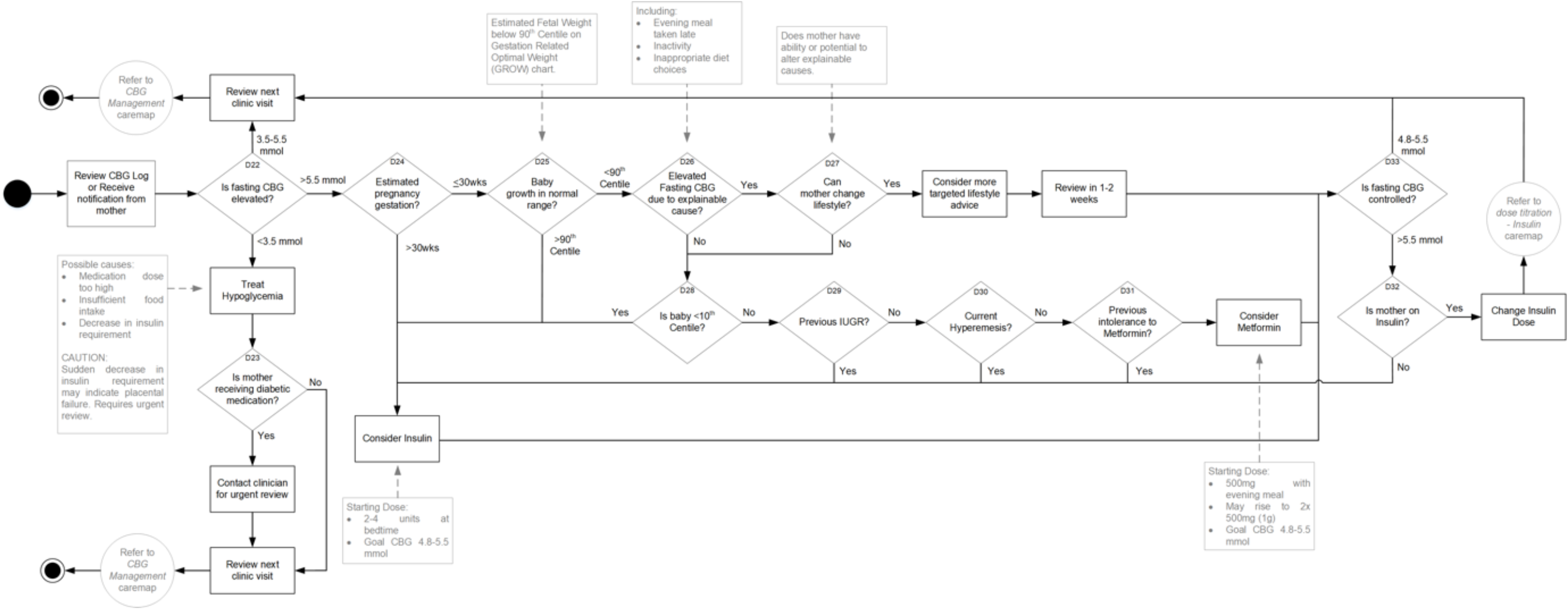
Treatment Decision for Fasting CBG Level

**Caremap 6.**
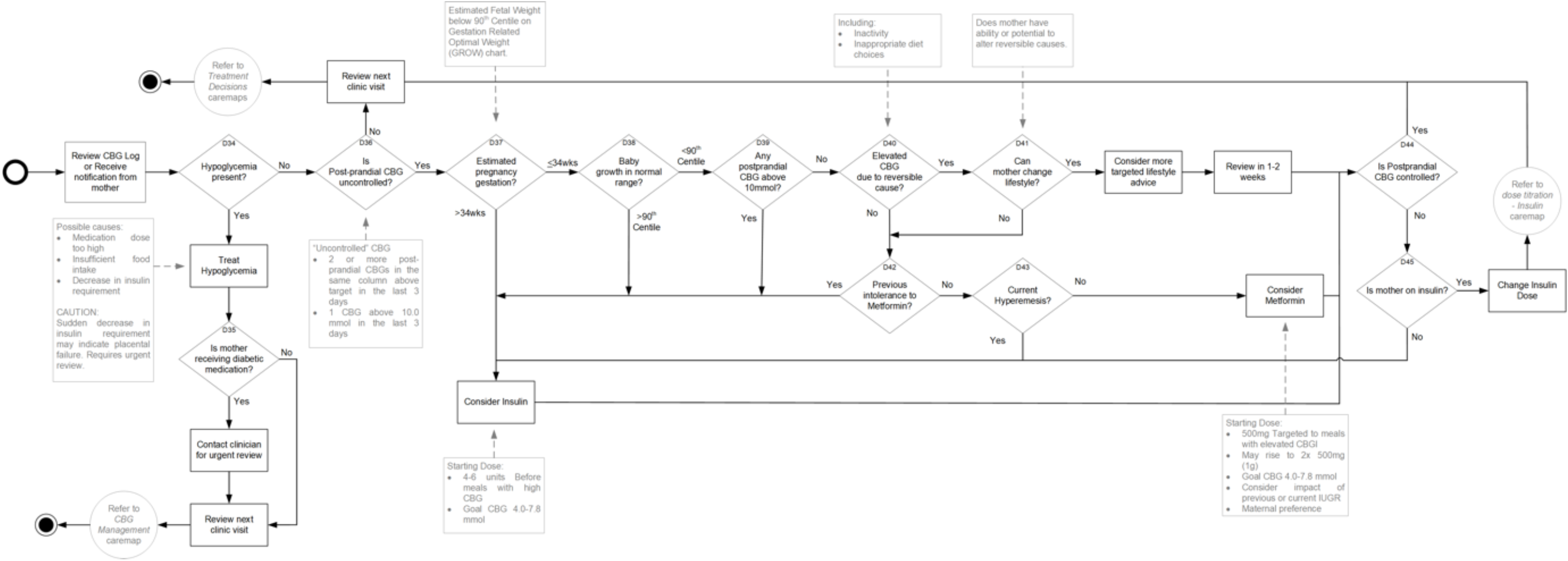
Treatment Decision for Post Prandial CBG Level

**Caremap 7.**
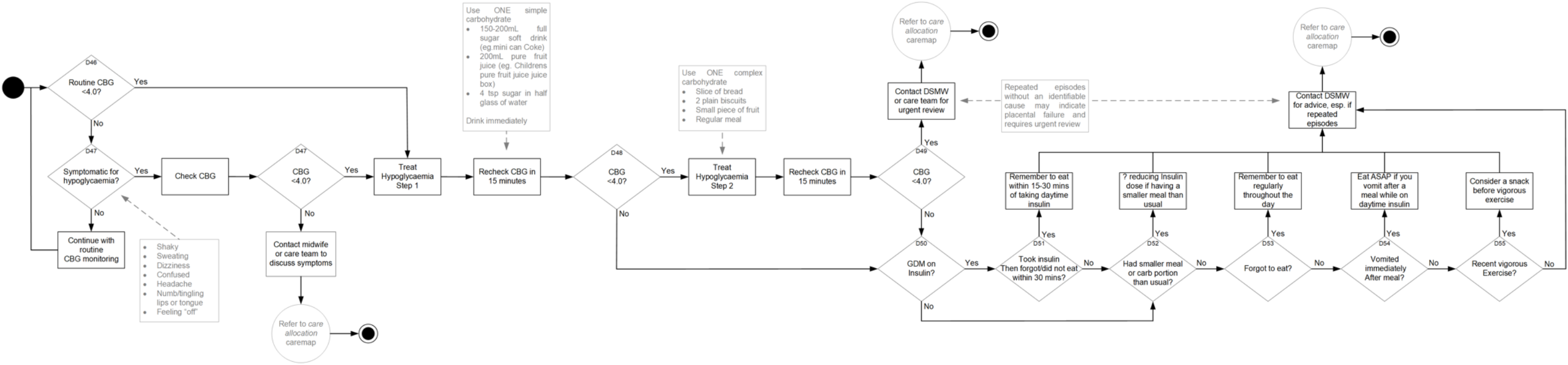
Hypoglycaemia

### 4.7 Diet Advice

While NICE recommends offering dietary advice (sections 1.2.14 - 1.2.15, 1.2.18, 1.6.11), it remains silent about the detail such advice should provide. How diet should be composed for women with GDM is a complex issue that remains largely unresolved [16]. While the primary focus for dietary recommendations is maintaining glucose levels within normal levels, a significant goal is also preventing excessive weight gain [17], which, in women with GDM is associated with an increased risk of: (i) hypertensive disorders of pregnancy including pre-eclampsia; (ii) caesarean section; and (iii) large for gestational age (LGA) babies. For these reasons the NICE guideline recommends referring the woman to a dietician (section 1.2.16). Creation of a caremap covering the dietetic involvement was outside the obstetric and midwifery remit of this work.

### 4.8 Dose Titration - Insulin

Caremap 8 describes the process for titrating dosage where the woman is receiving insulin therapy and her glucose levels remain uncontrolled.

**Caremap 8.**
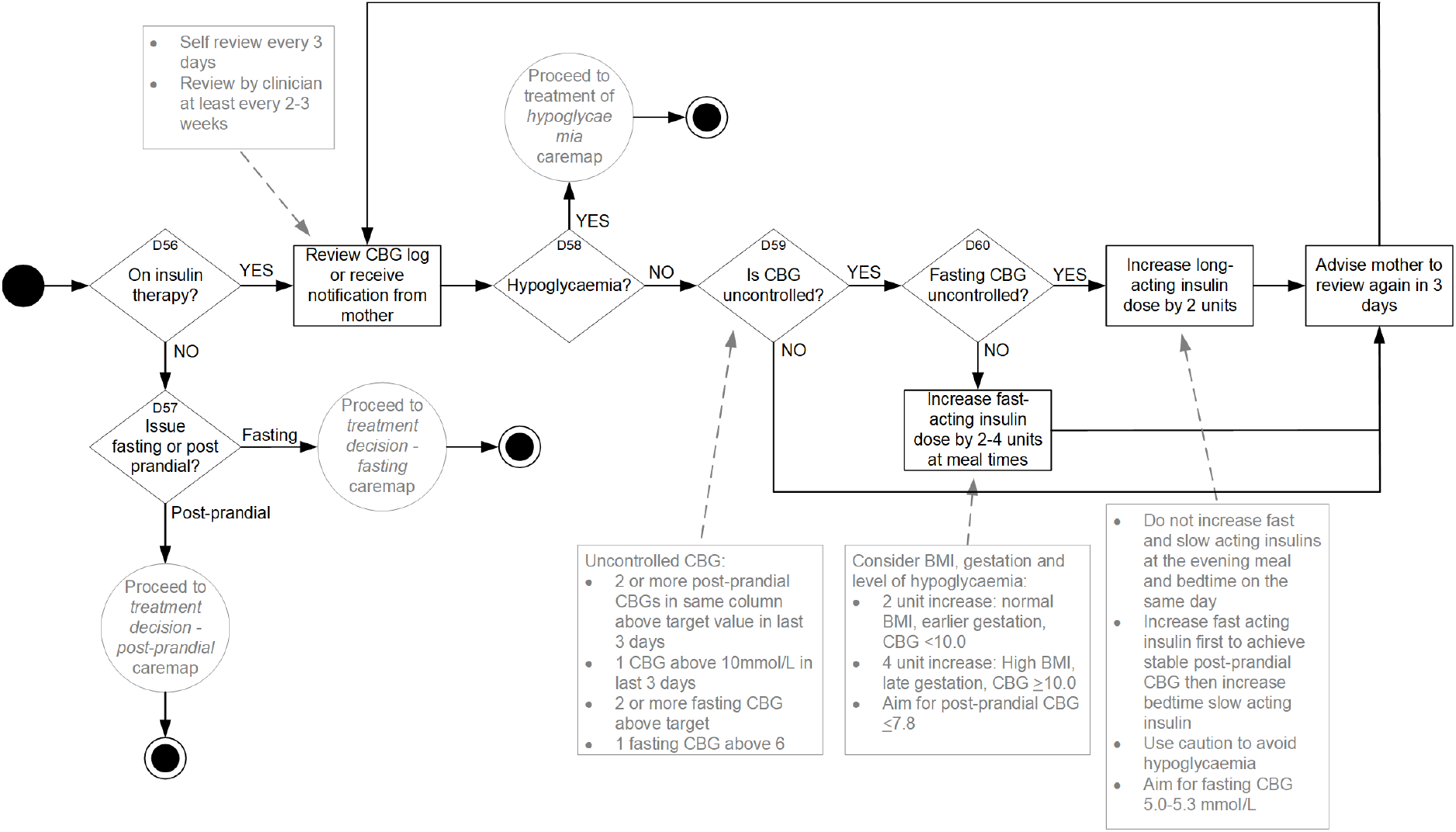
Dose titration of Insulin

### 4.9 Dose Titration - Metformin

Caremap 9 describes the process for titrating dosage where the woman is receiving Metformin therapy and her glucose levels remain uncontrolled.

**Caremap 9.**
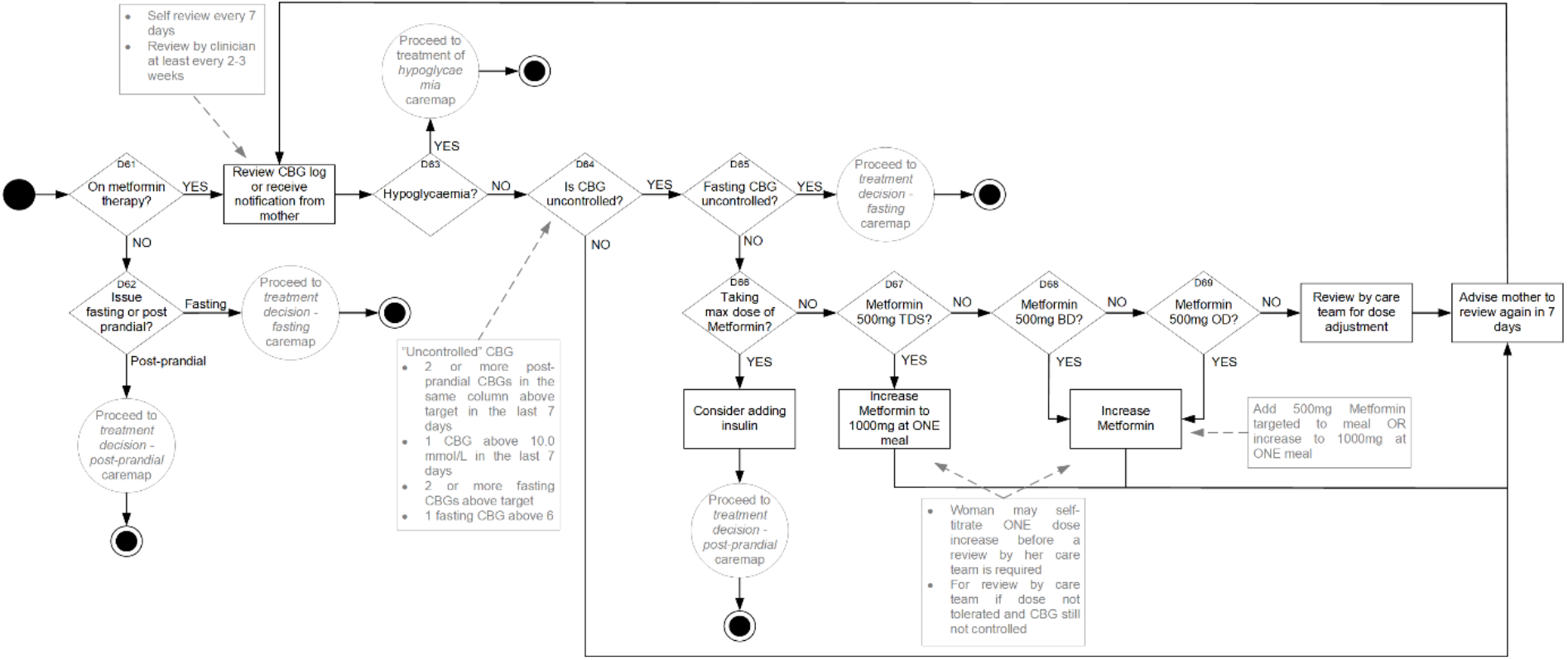
Dose titration of Metformin

### 4.10 Delivery Decision

This important caremap incorporates recommendations from sections 1.4.1 - 1.4.7 of the NICE guideline [15]. While an algorithmic approach can be described based on the guideline for the delivery decision, situations may remain that fall outside any such approach when the mother has other obstetric concerns and requires obstetric review prior to the delivery decision. While the final decision will be made in collaboration with the mother and her obstetric and midwifery teams, this decision is always subject to change if new information or complications are identified during labour.

**Caremap 10.**
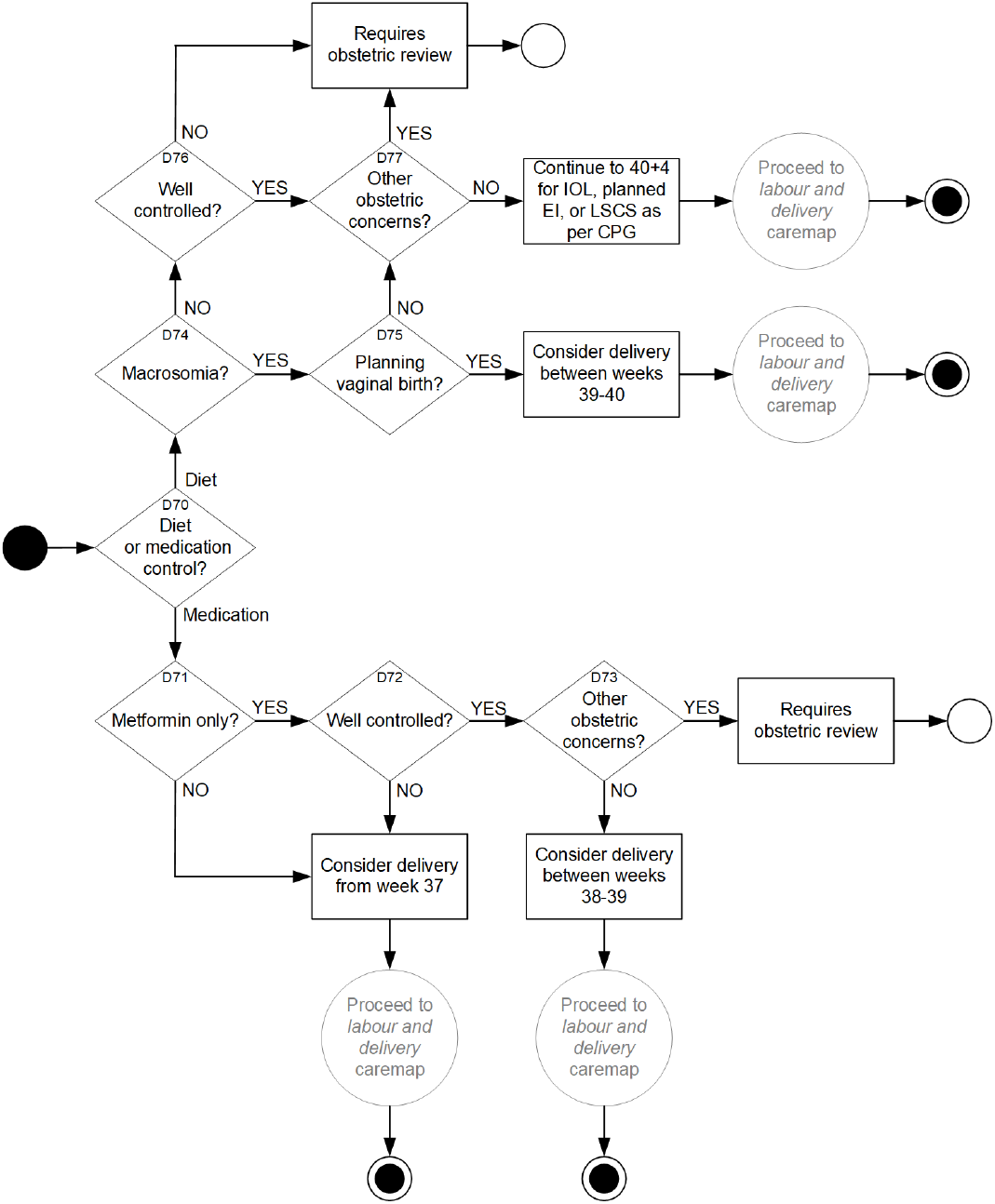
Delivery Decision

### 4.11 Labour and Delivery

The labour and delivery caremap commences when labour is established and ends after placental delivery. Development of our labour and birth caremap was first described in McLachlan [18]. That caremap comprehensively considered the possible pathways, interventions and outcomes (spontaneous vaginal delivery, emergency or elective caesarean, assisted delivery with forceps or ventous, spontaneous breech delivery, and rare cases of birth before arrival at the hospital). The labour and delivery caremap’s accuracy was evaluated in that work through critical review by midwives and obstetricians who were part of the research team, and as shown in the State Transition Machine (STM) reproduced from that work in Figure 3, by application of statistics covering all births for a single birthing centre for an entire year^1^ to demonstrate a contiguous pathway for care and outcome could be established for all pregnant and labouring women on entering the caremap [18].

**Figure 3:**
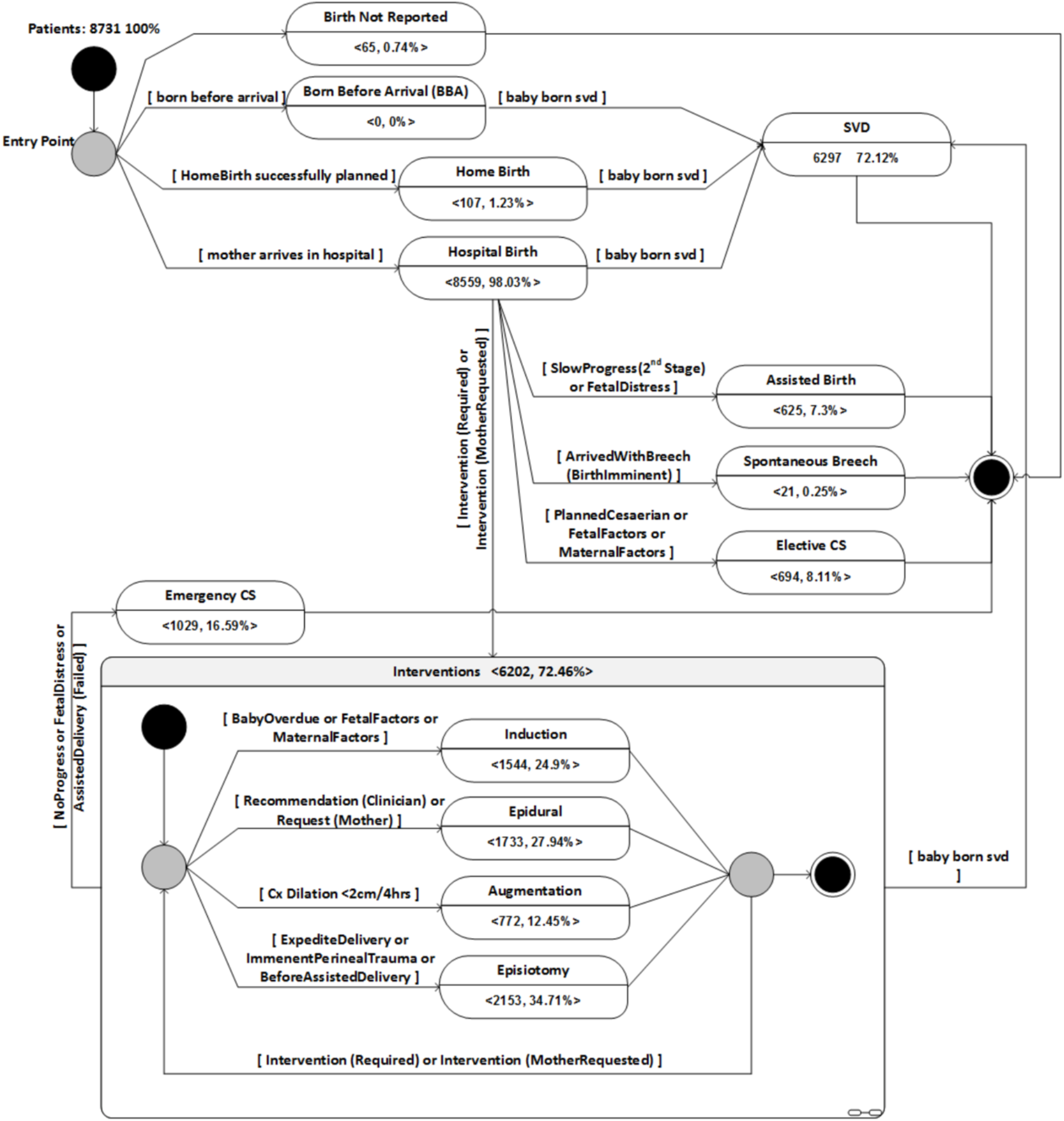
Labour and Birth caremap from McLachlan et al (2015) adapted as a State Transition Machine (STM) for use in development and training of artificial intelligence (AI) and synthetic data generation (SDG) incorporating patient outcome data describing all births in the Counties Manukau District Health Board catchment area of New Zealand for 2014.

### 4.12 Postnatal Testing

The *postnatal testing* caremap evaluates whether glucose control returns to normal prior to discharging the woman to primary care at 6 weeks postnatal (NICE guideline section 1.6.8). Women diagnosed with GDM are advised regarding the potential for GDM to return during future pregnancies (NICE guideline section 1.6.10). All women who have had GDM during their pregnancy are recommended to have their HbA1C tested at week 13 (NICE guideline section 1.2.11). Where the HbA1C test returns a result in the normal range they will be provided with additional lifestyle advice, informed of the potential risk for future T2DM, and recommended to have annual glucose testing to monitor for future diabetic disease (NICE guideline section 1.6.13). While her GDM status will always be provided with any discharge notes to the GP, where the woman returns abnormal glucose results to these tests she will also be advised this may indicate positive diagnosis for T2DM and offered referral to the local diabetic service of the healthcare provider (NICE guideline section 1.6.13).

**Caremap 11.**
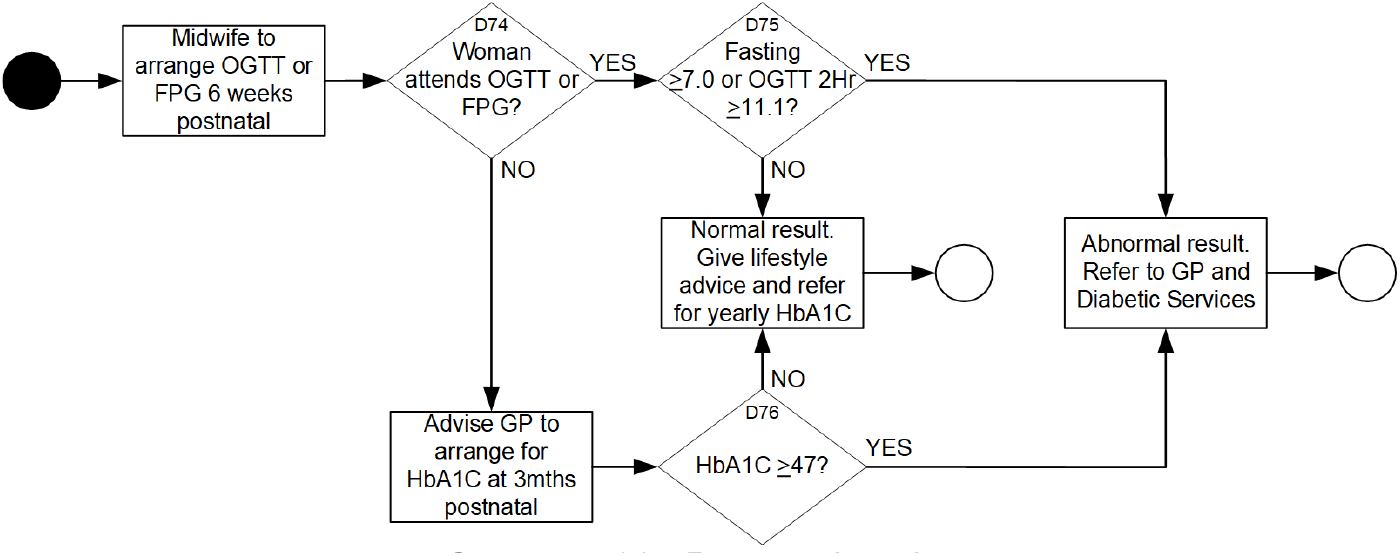
Postnatal testing

## 5 Using the HCT and Caremaps to develop clinical decision support tools

Once the GDM caremaps were verified, the research midwife commenced data collection from pregnant women with identified risk factors for GDM, whether or not they went on to develop and be diagnosed with the condition. A complex mix of qualitative and quantitative information supports modelling of the decision points identified in the GDM caremaps. Inclusion of data from patients with risk factors but both with and without the condition infuses the decision support tool with an understanding for what is, and is not, that condition. The second process stream followed steps iii, iv and v as described in the method. While the entire HCT consists of twelve caremaps and more than thirty primary clinical decision points, there is not always a one-to-one relationship between a caremap or decision point and a Bayesian network (BN). The number of required BNs depends on a range of factors, not limited to the types of decisions being modelled, who from and how input is being provided, and who will receive and how the output will be used. The rest of this section provides examples of the main steps for developing a single BN model for one caremap - the *care allocation* caremap.

BNs are probabilistic directed acyclic graphs with two components: The first is *qualitative*, or the structure, which comprises the *nodes* (corresponding to variables of interest) and *arcs* (edges) that indicate dependence between nodes. The second is *quantitative*, consisting of probability tables associated with each node (not shown). These probability tables contain the parameters (probabilities) associated with each state of the node. For nodes without parents these are the prior probabilities for each state, and for nodes with parents these are the conditional probabilities given the combined parent state. The probability parameters are generally determined from a combination of data and knowledge. Once the BN structure and probability tables are defined, it can be used for decision support by entering observed values for any known variable. Bayesian propagation calculates the revised probabilities of all unknown variables.

### Identification of Idioms

An introduction to the concept of idioms and their use in developing BNs can be found in [34] starting at page 204. Since their introduction, the idiomatic approach for developing structurally explainable BNs has been extended to describe common reasoning patterns specific to the domains of law [35] and medicine [36].

### Care allocation

We identified an instance of the *definitional/synthesis idiom* described in [34] at 8.3.3 explaining one of the reasoning processes of the clinical decision-making task described by our clinical experts and expressed in Caremaps 1 and 2. The *definitional/synthesis idiom* combines identified variables into a sensible and easy-to-read hierarchical structure using synthetic nodes to bring together parent nodes that are factors, or components, of common concepts. The medical version of the definitional/synthesis idiom shown in Figure 4b was described in [36] and illustrates the hierarchical nature and use of synthetic nodes through the *Medical History* example - in that while the *Comorbid Disease* and a *Family History of Disease X* parent nodes could be connected directly to the *Likelihood of Disease X* child node, they are both factors common to *Medical History* and thus are easier to visualise, explain, and understand when the *Medical History* synthetic node is present.

**Figure 4:**
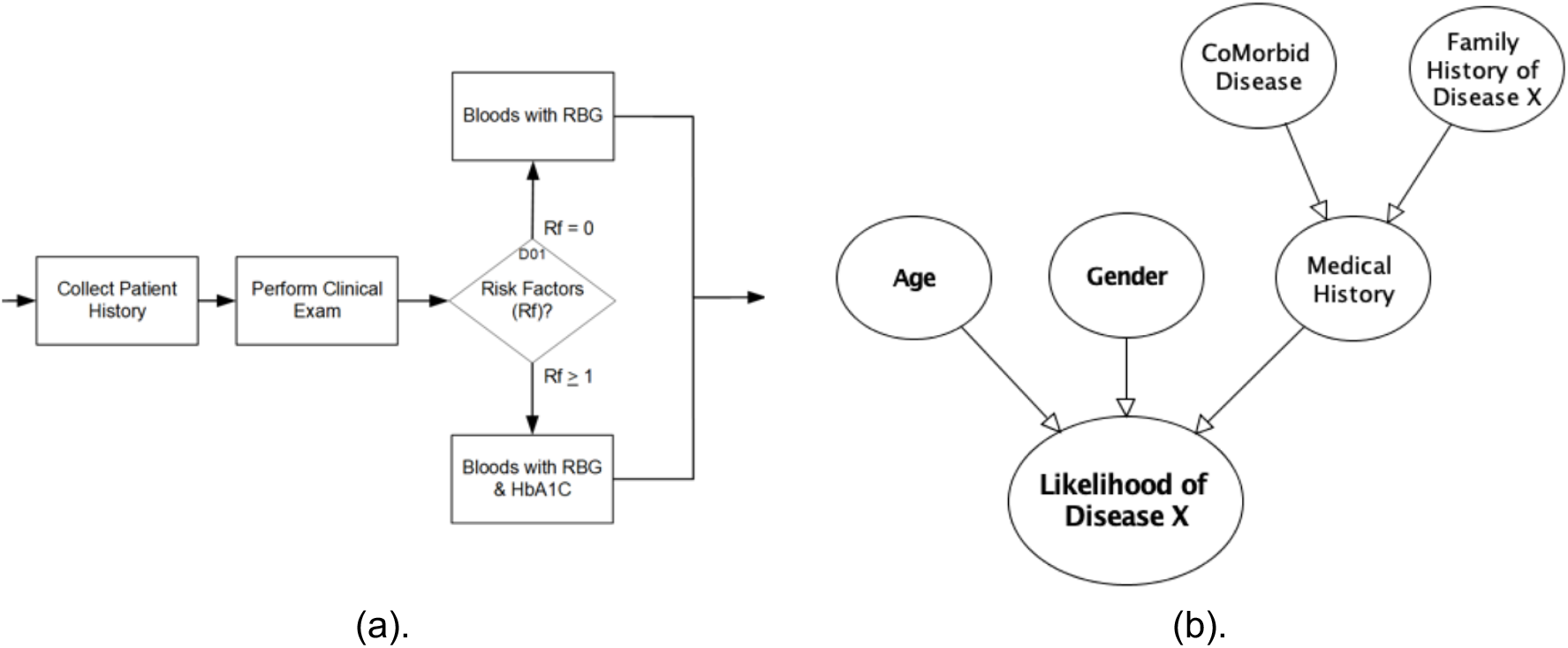
(a) Fragment of Booking Visit caremap showing where patient medical history and risk factors are collected also showing a decision point regarding the blood tests required in which the clinician evaluates risk factors that predispose the likelihood that the patient has the disease; (b) Medical version of definitional/synthesis idiom from Kyrimi et al 2020 that describes the structure of clinical reasoning for decision-making regarding the likelihood that the patient has the disease.

The BN structure fragment shown in Figure 5 evaluates observations to decide whether the woman is at an elevated obstetric risk. Obstetric risk is computed as the combined effect of observing the presence or absence of known GDM risk factors that include current pregnancy complications, past obstetric concerns, comorbid medical conditions and the pregnant woman’s BMI.

**Figure 5:**
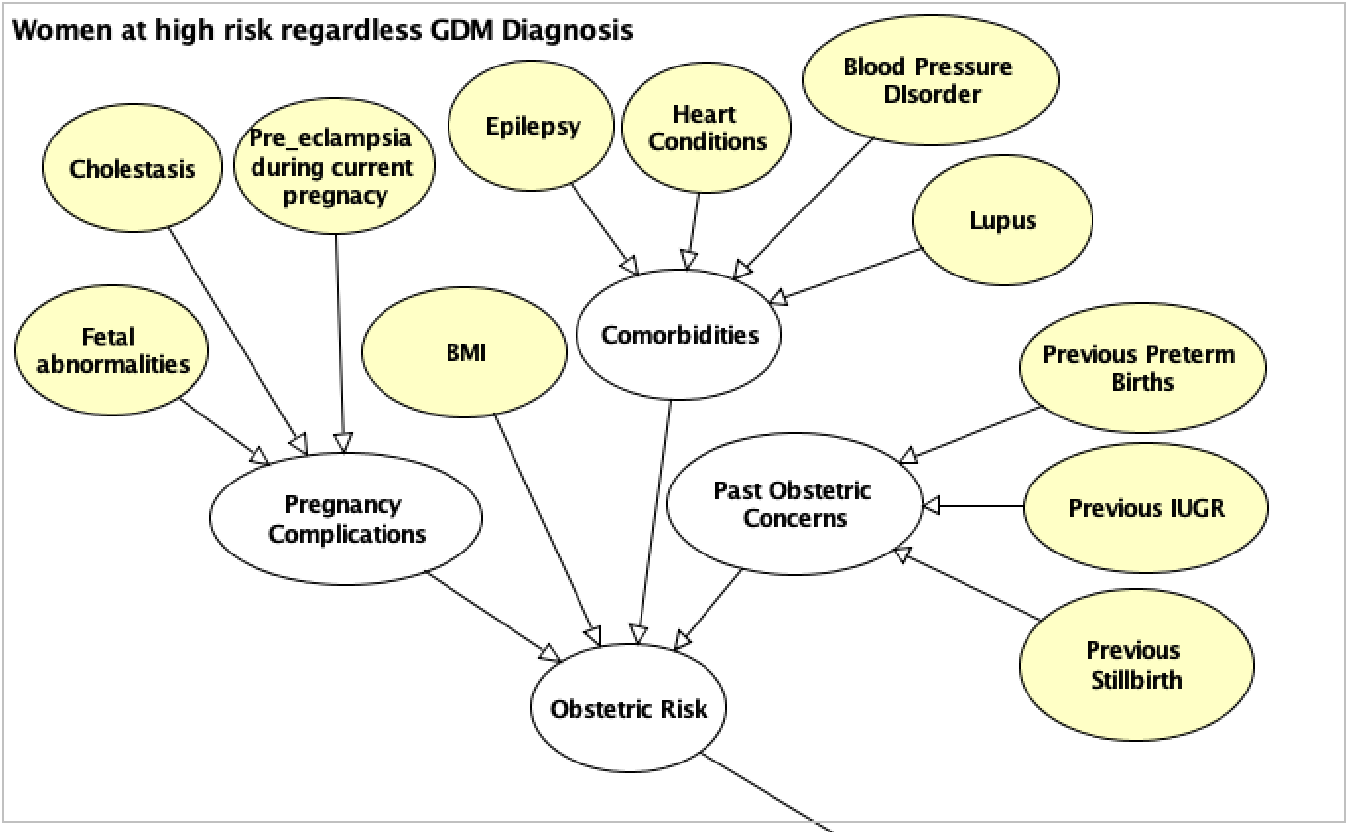
Application of the Medical Definitional/Synthesis Idiom

### Identification of Variables and Parameters

Variables were identified as an output of the process leading from expert elicitation through caremap modelling to identification of idioms. In consultation with our clinical experts, the origin (if retrospective) or source (if prospective), instantiation, data entry or data collection time and functional notes were recorded. Table 1 shows an extract of the variable description table.

**Table 1:**
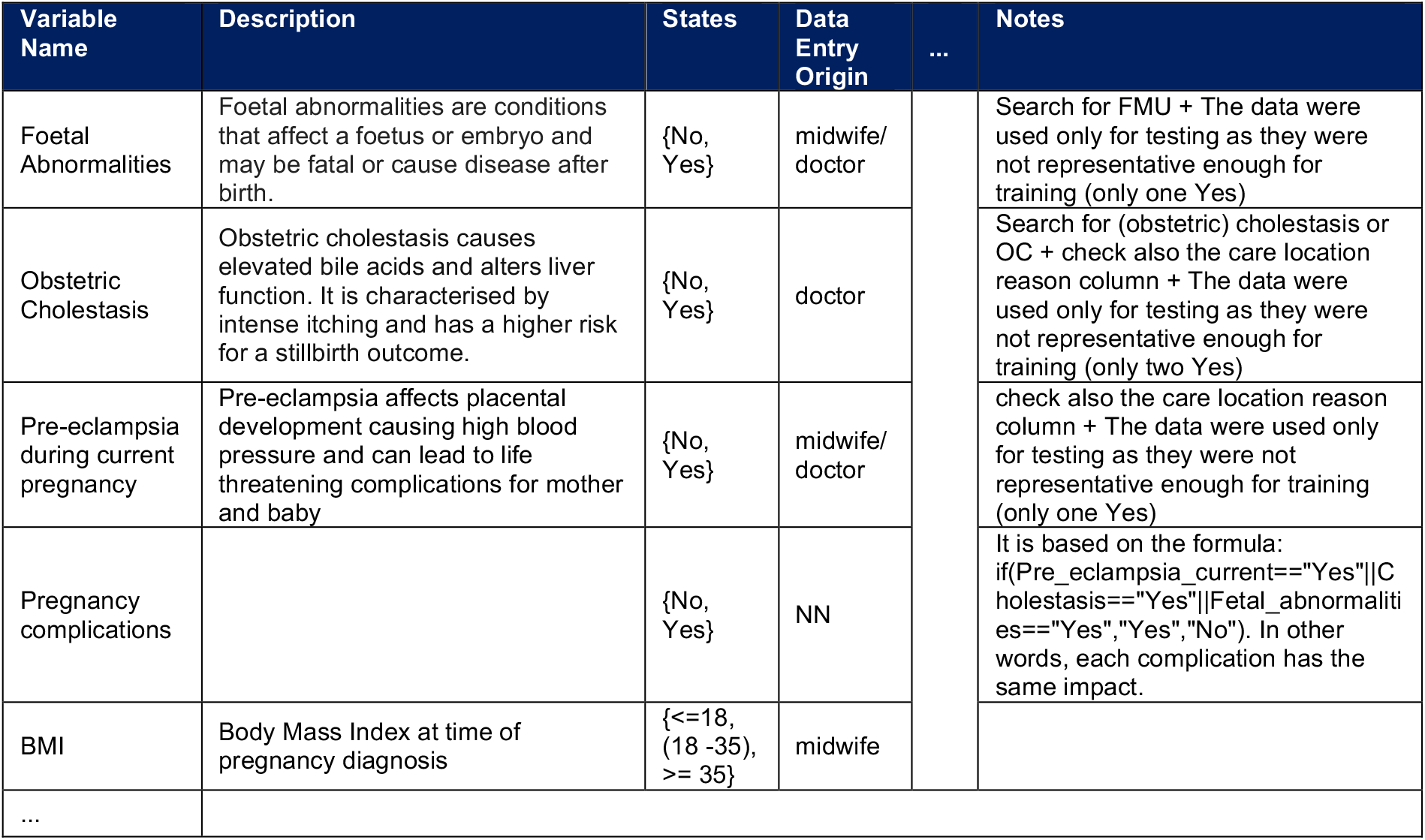
Variable Description Table

The probability values of variables in medical BNs can be learned or observed, either *retrospectively* or *prospectively*. Retrospectively when identified from the patient’s medical history, or prospectively when acquired in real-time (or near real-time) from a monitoring device such as a capillary blood glucose (CPG) meter, or when the patient enters them into the model via a form or ‘app’.

The research midwife (RM) was responsible for data collection from EHR (retrospective) or three times per week from in-patient treatment files (prospective). To protect patient privacy and meet ethics requirements, each patient was reported via a randomly generated *subject ID*. The RM maintained a separate linking table on her hospital computer to assist when data analysts and decision scientists asked questions regarding a particular patient record. The RM could investigate and respond to their requests. Other data were also anonymised, including replacing the *date of birth* with an age in years, and delta shifting all dates in the patient record using a consistent randomly assigned number of days for each patient. All examples provided in the tables in this work are representative of the real anonymised data collected and used during the project, but were synthetically generated for this paper using the method described in [37]. Table 2 shows an example of raw patient demographics data.

**Table 2:** Raw Data - Patient Demographics

| Subj_ID | Pregnancy Complications | Care Location Reason | BMI | Medical Conditions | ... | Care Location |
| --- | --- | --- | --- | --- | --- | --- |
| NLM1803 | PRE DIABETES IN 2018 (LAST PREGNANCY) | HIGH BMI, HBA1C AT BOOKING | 42.97 | NIL |  | DANC |
| BBC0999 | RAISED HBA1C AT BOOKING, GLUCOSURIA ++ IN CLINIC | PREV GDM, PREV IUGR, RAISED HBA1C AT BOOKING | 30.80 | NIL |  | DANC |
| PYK9441 | OLIGOHYDRAMNIOS | PREV IUGR, PRE ECLAMPSIA, PRE TERM | 29.58 | NIL |  | DANC |
| NGU2893 | ERRACTIC CBGS, NOT CHECKING AT CORRECT TIMES | HIGH BMI, HBA1C AT BOOKING | 39.33 | NIL |  | DANC |
| ZBR9059 | NIL | HIGH BMI, HBA1C AT BOOKING | 43.12 | FGM |  | DANC |
| ZJY4761 | POOR RECORDER OF CBGS | NEEDS TREATMENT, POOR COMPLIANCE | 28.46 | NIL |  | COMMUNITY |
| ... |  |  |  |  |  |  |

Women with GDM are required to self-perform capillary blood glucose (CBG) tests up to four times each day - prior to breakfast (fasting), after breakfast (AB), after lunch (AL) and after the evening meal (AE). Table 3 shows a synthetically generated example of raw CBG data.

**Table 3:** Raw Data - Capillary Blood Glucose

| Subj_ID | Date | Fasting | AB | AL | AE |
| --- | --- | --- | --- | --- | --- |
| DNJ8354 | 20/12/2018 | 5.2 | 5.8 | 6.0 | 5.0 |
| DNJ8354 | 21/12/2018 | 5.8 | 4.9 | 5.9 | 5.0 |
| DNJ8354 | 22/12/2018 | 5.4 | 5.1 | 5.8 | 4.9 |
| DNJ8354 | 23/12/2018 | 5.5 | 5.1 | 7.9 | 4.9 |
| DNJ8354 | 24/12/2018 | 5.4 | 5.2 | 7.9 | 4.8 |
| DNJ8354 | 25/12/2018 | 5.2 | 5.3 | 8.1 | 4.7 |

### Discretising and Coding

Raw data for populating conditional probability tables (CPT) of a variable (node) can present with continuous or discrete values. All continuous variables (such as age, BMI) have been mapped into discrete scales. Table 4 shows the output of discretising the patient demographics data from Table 2.

**Table 4:** Prepared Data - Patient Demographics

| Subj_ID | Foetal Abnormalities | Cholestasis | Pre Eclampsia current | BMI | ... | Care Location |
| --- | --- | --- | --- | --- | --- | --- |
| NLM1803 | No | No | No | >=35 |  | DANC |
| BBC0999 | No | No | No | 18-35 |  | DANC |
| PYK9441 | No | No | No | 18-35 |  | DANC |
| NGU2893 | No | No | No | >=35 |  | DANC |
| ZBR9059 | No | No | No | >=35 |  | DANC |
| ZJY4761 | No | No | No | 18-35 |  | COMMUNITY |
| ... |  |  |  |  |  |  |

The CBG data was collected as four readings per day, almost every day for up to eight months. While values for each measurement ranged from 0.2 to 26.7 millimoles per litre (mmol/L), this degree of granularity was unnecessary. When three days measurements with four measurements per day would be processed in any operation of the BN, this complexity would have significantly increased processing overhead. The CBG data was discretised to diagnostic thresholds set by the CPG of the treating hospital, which were:

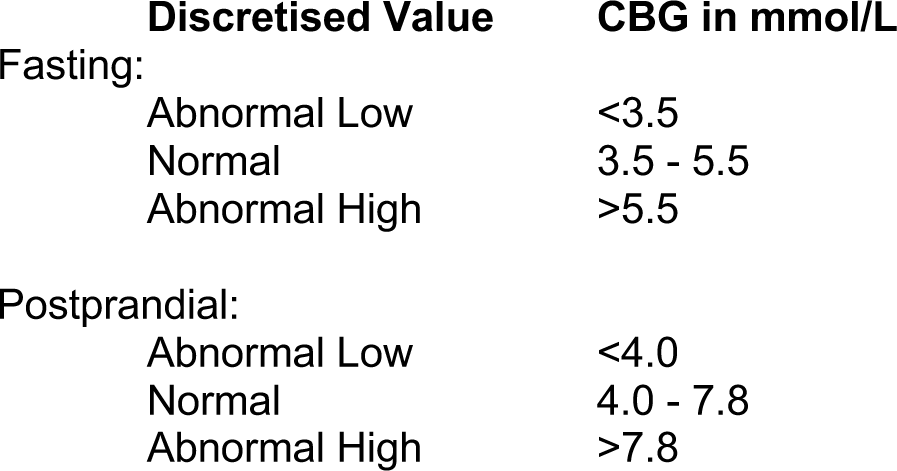

Table 5 shows discretised CBG data. The current day’s data is labelled with a T (current time), while data for each of the two previous days is labelled as T1 (time 1 day previous) and T2 (time 2 days previous).

**Table 5:** Prepared Data - Capillary Blood Glucose

| Subj_ID | Date | Fasting<br>T | Fasting<br>T 1 | Fasting<br>T 2 | AB<br>T | AB<br>T 1 | AB<br>T 2 | AL<br>T | ... |
| --- | --- | --- | --- | --- | --- | --- | --- | --- | --- |
| DNJ8354 | 20/12/2018 | Normal | Normal | Normal | Normal | Normal | Normal | Normal |  |
| DNJ8354 | 21/12/2018 | Abnormal<br>H | Normal | Normal | Normal | Normal | Normal | Normal |  |
| DNJ8354 | 22/12/2018 | Normal | Abnormal<br>H | Normal | Normal | Normal | Normal | Normal |  |
| DNJ8354 | 23/12/2018 | Normal | Normal | Abnormal<br>H | Normal | Normal | Normal | Abnormal<br>H |  |
| DNJ8354 | 24/12/2018 | Normal | Normal | Normal | Normal | Normal | Normal | Abnormal<br>H |  |
| DNJ8354 | 25/12/2018 | Normal | Normal | Normal | Normal | Normal | Normal | Abnormal<br>H |  |

### The Care Allocation BN

To support clinical decision-making, the resulting *care allocation* BN must emulate clinical expert reasoning. However, while the clinician may become less accurate where information is absent, such as test result values or elements of the patient’s medical history, a key benefit of BNs is their ability to reason with missing data.

The full *care allocation* BN shown in Figure 6 factors:

**Figure 6:**
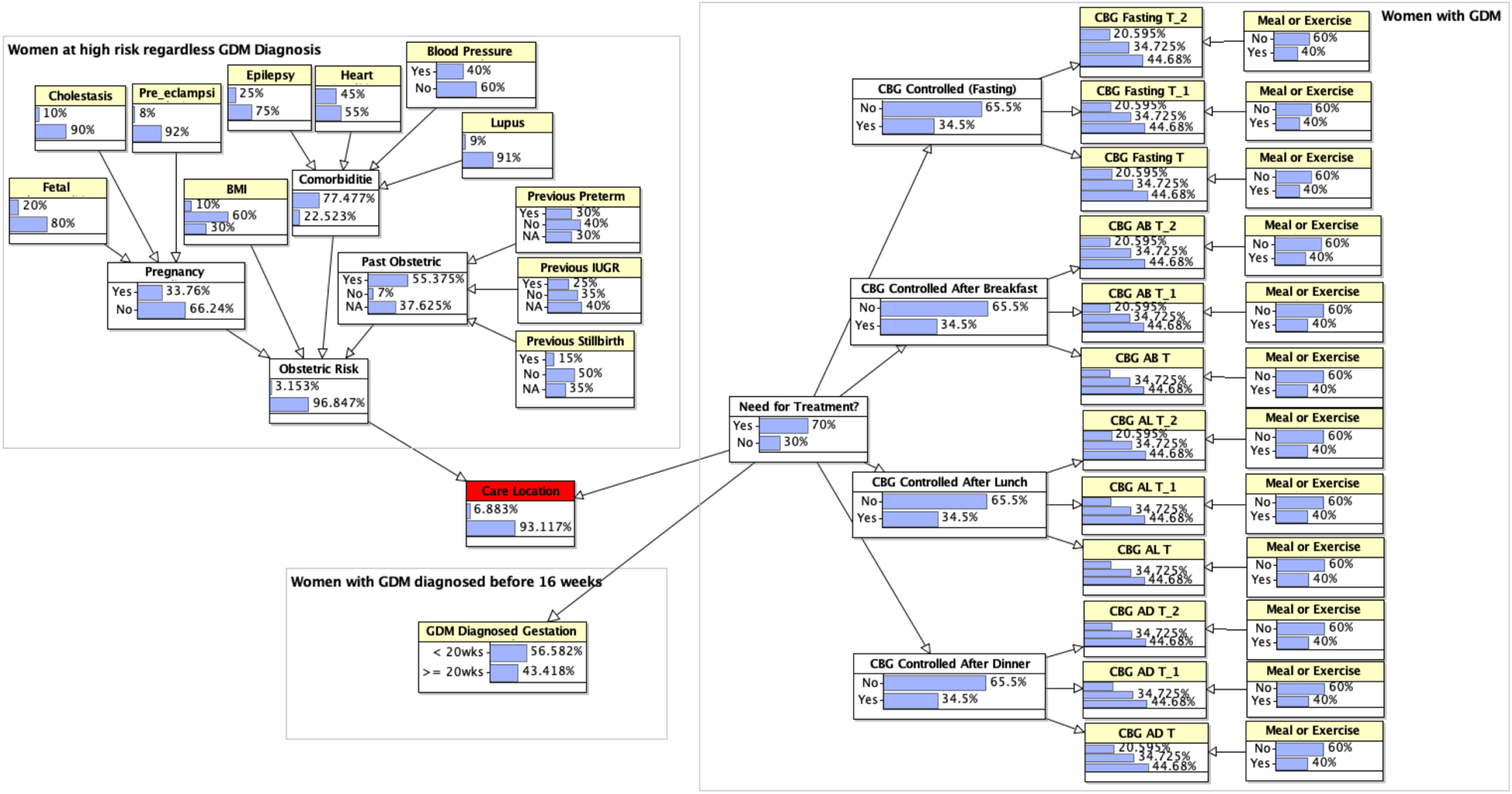
Full Bayesian Network - Care Allocation

1. The level of obstetric risk based on relevant factors from the woman’s current health and physical state, past pregnancies, and overall medical history;
2. Whether the woman’s GDM was diagnosed before or after 16 weeks gestation; and,
3. The current CBG result with the two previous day’s CBG;

The BN was trained using retrospective data from 250 patients with risk factors for GDM, whether or not they went on to be diagnosed with the disease.

Figure 6 also shows the marginal probabilities for each node (i.e. this represents what we know about the whole population before observations specific to the individual is entered). The model was validated with a prospective dataset containing 124 patients who were cared for by 2 diabetologists, 3 obstetricians and a team of 8 community midwives (13 clinical staff in total), and who had delivered prior to completion of this portion of the project. The model provided care allocation decisions that were consistent with those of the clinician for 116 of the 124 prospective patients. Analysis of the 8 cases where outcomes were in disagreement found them to be examples of the application of clinical judgement over blind adherence to guidelines. In each case the clinician had either acted pre-emptively to escalate referral for a woman they believed would soon develop GDM, or had elected to wait and see for a woman whose condition barely met the diagnostic criteria.

## 6. Next steps and future work

The project also produced: a prototype BN for managing glucose control that, with further specification, may also titrate metformin dosage [38]; and an architecture and prototype smartphone and web-based application for use by clinicians and patients [33]. However, the tasks of creating a full end-to-end suite of BNs covering all necessary decision points in the GDM HCT and clinical trials remain. When the entire suite of BN models are complete and integrated into the decision support application, we suggest a more formal method that easily maps and integrates the HCT and caremaps with decision technologies, especially BNs, and a side-by-side outcome validation of the application’s decisions through comparison to those of clinicians at each step during the patient management process. The proposed approach would allow testing and validation of the models and application during entire pregnancies without affecting clinical care, as clinicians and midwives would continue to make decisions for their pregnant women with no visibility of the application’s output. It will also allow us to see how well the application updates its own decision models as it receives new evidence regarding the pregnant woman’s ongoing health and GDM condition. It has been suggested that the completed GDM tool could be beneficial to those in remote regions of countries like Zimbabwe where many diabetic people, including pregnant women, die needlessly early because they receive little or no healthcare for their condition.

## 7. Conclusion

Like their component caremaps, HCT identify task or activity sequences and represent them using consistent visual features. For this reason, and while they were originally described within the limited role of birth-to-death health and disease timeline visualisation, HCT may be appropriate for a broad range of tasks not limited to: care quality improvement, clinical training and patient education, systems and solution development, and specification of a wide range of research tasks. In particular, we propose the GDM HCT could be used in: (i) obstetric, midwife and patient education as a tool to describe the timeline, treatment pathways, options and potential outcomes for pregnant women diagnosed with GDM; and (ii) development of clinician- and patient-facing software applications to monitor the gestationally diabetic woman’s condition and support real-time clinical decision-making in both community (Primary) and clinical (secondary and tertiary) settings. While caremaps have received renewed attention during the last five years, and their representative language has been refined and standardised, as with most process flow diagrams in medicine until now they have remained limited to a single clinical incident or care event. The HCT method had been previously proposed for creation of a health and disease lifecycle, however it had not been used to model an entire disease lifecycle from diagnosis to cure or, if chronic, ongoing management. Furthermore, hitherto, little or no effort has gone into investigating methods for seamlessly integrating the HCT and caremaps with decision models such as Bayesian Networks as presented in this paper. HCT have applications in clinical quality improvement, management, education and care, and could be used as an aid when explaining complex medical conditions with multiple treatment pathways to patients. We have also shown that caremaps can be used to expedite development of artificial intelligence for clinical decision-support and clinician- and patient-facing applications. By creating Bayesian models to support all of the main decision points of the entire HCT, an end-to-end tool could be developed that would enable patient self-management. This could improve patient awareness for their condition and reduce the impact of their disease on themselves and the limited resources of our healthcare systems.

## Data Availability

No Data is necessary

## Glossary of Terms

BN: Bayesian network
CBG: Capillary Blood Glucose
CPG: Clinical Practice Guideline
CPT: Conditional Probability Table
DSMW: Diabetic Specialist Midwife
EHR: Electronic Health Record
EPSRC: Engineering and Physical Sciences Research Council
GDM: Gestational Diabetes Mellitus
HCT: Health Condition Timeline
NICE: National Institute for Health and Care Excellence
OGTT: Oral Glucose Tolerance Test
PBG: Plasma Blood Glucose
RS-EHR: Realistic Synthetic Electronic Health Records
RS-HCT: Realistic Synthetic Health Condition Timeline

## Ethics

The regular Clinical Audit of the Diabetes in Pregnancy (gestational diabetes) pathway that was used to produce anonymised aggregate statistics to train and validate the Bayesian model was approved by the Barts Health NHS Trust. 14/12/2018. Audit ID: 9861 and 08/05/2017. Audit ID: 8268. Requirements for further ERB/IRB or patient consent were waived.

## Footnotes

1 In the example shown, Middlemore Hospital of the Counties Manukau District Health Board in New Zealand during 2014.

